# Association of Baseline Cerebrovascular Reactivity and Longitudinal Development of Enlarged Perivascular Spaces in the Basal Ganglia

**DOI:** 10.1101/2023.05.14.23289968

**Authors:** T.J. Libecap, Christopher E. Bauer, Valentinos Zachariou, Colleen A. Pappas, Flavius D. Raslau, Peiying Liu, Hanzhang Lu, Brian T. Gold

**Affiliations:** MD/PhD Program, University of Kentucky College of Medicine, Lexington, Kentucky, USA; Department of Neuroscience, University of Kentucky College of Medicine, Lexington, Kentucky, USA; Department of Radiology, University of Kentucky College of Medicine, Lexington, Kentucky, USA; Department of Radiology, University of Maryland School of Medicine, Baltimore, Maryland, USA; Department of Radiology, Johns Hopkins University School of Medicine, Baltimore, Maryland, USA; Magnetic Resonance Imaging and Spectroscopy Center, University of Kentucky, Lexington, Kentucky, USA; Sanders-Brown Center on Aging University of Kentucky, Lexington, Kentucky, USA

## Abstract

1.

**Background:** Increasing evidence suggests that enlarged perivascular spaces (ePVS) are associated with cognitive dysfunction in aging. However, the etiology of ePVS remains unknown. Here we tested the possibility that baseline cerebrovascular dysfunction, as measured by an MRI measure of cerebrovascular reactivity (CVR), contributes to the later development of ePVS.

**Methods:** A total of 79 cognitively normal, older adults (46 women, age range 60-84) were recruited to undergo MRI scanning at baseline and 50 participants returned for a follow-up scan approximately 2.5 years later. ePVS were counted in the basal ganglia, centrum semiovale, midbrain, and hippocampus. CVR, an index of the vasodilatory capacity of cerebral small vessels, was assessed using carbon-dioxide inhalation while acquiring blood oxygen-level dependent (BOLD) MR images.

**Results:** Low baseline CVR values in the basal ganglia were associated with increased follow-up ePVS counts in the basal ganglia after controlling for age, sex, and baseline ePVS values (coefficient estimate (SE) = -15.87 (3.92), p < 0.001, 95% confidence interval [CI] -23.68 to - 8.05). This effect remained significant after accounting for self-reported risk factors of cerebral small vessel disease (cSVD) (coefficient estimate (SE) = -15.03 (4.00), p < 0.001, CI -23.02 to - 7.05) and neuroimaging markers of cSVD (coefficient estimate (SE) = -13.99 (4.02), p < 0.001, CI -22.03 to -5.95).

**Conclusions:** Our results demonstrate that low baseline CVR is a risk factor for later development of ePVS. MRI-based CVR may represent a promising biomarker of cSVD.

## 2. Introduction

Perivascular spaces (PVS) are pial-lined, fluid-filled spaces surrounding penetrating brain arteries ^1^. PVS are integral to neuroimmune function and clearance of metabolites via the brain’s glymphatic system as the site of interchange between interstitial fluid (ISF) and cerebrospinal fluid (CSF) ^2,3^. Enlarged perivascular spaces (ePVS) can be detected as dilated PVS on magnetic resonance imaging (MRI). While originally considered a benign radiological finding, increasing evidence suggests that ePVS may represent microvascular dysfunction and impaired ISF-CSF interchange ^1,4^.

Consistent with this possibility, a body of work has shown ePVS are associated with cognitive dysfunction in aging ^4–8^. For example, we recently found that greater ePVS counts are associated with lower scores on the Montreal Cognitive Assessment (MoCA), a standardized clinical tool of global cognitive performance ^8^. In addition, two recent studies showed a relationship between high ePVS burden and worse executive function performance ^6,7^.

Despite such evidence supporting the clinical significance of ePVS, mechanistic contributors to ePVS remain under-characterized. Functionally-based cerebrovascular dysfunction could represent a contributing factor to structural PVS enlargement. For instance, reduced vascular compliance could disrupt ISF-CSF interchange and impair waste clearance ^4,9^. The reduced clearance of brain waste could in turn promote PVS enlargement ^3^.

Testing this possibility requires a valid measure of cerebrovascular compliance. Recently, a blood oxygen-level dependent (BOLD)-fMRI cerebrovascular reactivity (CVR) method has been developed and validated as a measure of cerebrovascular compliance ^10–12^. This CVR method represents a functional assessment of the ability of the cerebrovascular system to dynamically respond to a vasodilatory stimulus ^11^. Specifically, participants’ cerebrovascular response to an in-scanner hypercapnia challenge is measured by dividing the percent change in BOLD-fMRI signal by the change in end-tidal CO_2_ (mmHg) between normal room air breathing and hypercapnic air breathing.

Importantly, several recent studies have described the cross-sectional association between CVR and ePVS ^13,14^, further motivating a hypothesis that low baseline CVR may contribute to later ePVS. Here, we tested this possibility in a longitudinal design involving a 2.5-year follow-up sample of older adults who were cognitively normal at baseline. Since the basal ganglia is a known predilection site of early cerebral small vessel disease (cSVD) ^15,16^ that is closely associated with cerebrovascular dysfunction ^17–19^, we hypothesized that low baseline CVR in the basal ganglia may predict increased ePVS counts in this region.

## 3. Materials and Methods

### 3.1 Participants

Eighty-one cognitively normal, older adults (age range 60-84, 46 women) were initially recruited for this longitudinal experiment. Participants were recruited from an existing longitudinal cohort at the Sanders-Brown Center on Aging (SBCoA) and the Lexington, KY community. Participants were informed at the time of enrollment that this was a longitudinal study which involved baseline MRI scanning and a follow-up MRI scan approximately 2.5 years later. All participants provided informed written consent at each study time point under a protocol approved by the Institutional Review Board of the University of Kentucky and this study is compliant with STROBE (Strengthening the Reporting of Observational Studies in Epidemiology) guidelines ^20^. Participants were asked not to drink caffeine-based products on the day of their scan to minimize potential confounding effects on CVR. All participants were cognitively unimpaired at baseline based on either 1) clinical consensus diagnosis and scores from the Uniform Data Set (UDS3) used by US ADCs (procedure outlined elsewhere ^21,22^) or 2) a score of 26 or higher on the Montreal Cognitive Assessment (MoCA) ^23^ for those participants recruited from the community.

Exclusion criteria at each time point were significant head injury (defined as loss of consciousness for more than 5 min), stroke, neurological disorders (e.g. epilepsy, Alzheimer’s disease) or major psychiatric disorders (e.g. schizophrenia, active clinical depression), claustrophobia, pacemakers, the presence of metal fragments or implants that are incompatible with MRI, or significant brain abnormalities detected during imaging. A board-certified neuroradiologist (F.D.R.) evaluated the T1-weighted (T1W) and fluid-attenuated inversion recovery (FLAIR) images for evidence of stroke or other clinically relevant abnormalities. This resulted in exclusion of two participants due to evidence of previous stroke (one participant) and hydrocephalus (one participant), not known at the time of enrollment. Participants were eligible for rescan regardless of their cognitive status at the follow-up scan. Detailed characteristics of the final group of 79 baseline participants and 50 follow-up participants are shown in Table 1.

**Table 1.** Demographics and Mean Cognitive Measures

|  | Baseline | Follow-up |
| --- | --- | --- |
| N | 79 | 50 |
| Sex (F:M) | 46:33 | 31:19 |
| Age Range (years) | 60-84 | 62-86 |
| Age (years) | 69.81 (5.34) | 72.6 (5.77) |
| Education (years) | 16.42 (2.48) | 16.82 (2.49) |
| Time Between Scans (years) | - | 2.557 (0.158) |
| MoCA | 26.62 (2.55) | 26.33 (2.40) |
| BMI | 28.14 (6.17) | 27.85 (6.28) |
| Hypertension, N (%) | 34 (43.0) | 20 (40.0) |
| Type 2 Diabetes, N (%) | 9 (11.4) | 5 (10.0) |
| Lacunes, N (%) | 25 (31.6) | 27 (54) |
| Cerebral microbleeds, N (%) | 35 (44.3) | 33 (66) |

The table lists the mean ( ± sd) for age, the female to male ratio, years of education, time between baseline and follow-up scan, MoCA scores, and BMI. The number and percentage, N (%), of participants positive for additional self-reported vascular risk factors and cSVD neuroimaging markers are also reported. Baseline MoCA scores were unavailable for 2 participants.

### 3.2 Magnetic Resonance Imaging Protocol

Participants were scanned in a Siemens 3T Prisma scanner (software version E11C), using a 64-channel head coil, at the University of Kentucky’s Magnetic Resonance Imaging and Spectroscopy Center (MRISC). The following scans were acquired: (1) a 3D multi-echo, T1-weighted magnetization prepared rapid gradient echo (ME-MPRAGE) scan, (2) a 3D fluid-attenuated inversion recovery (FLAIR) scan, (3) a 3D, multi-echo gradient-recalled echo scan used for quantitative susceptibility mapping (QSM), and (4) a BOLD EPI/fMRI scan for assessment of cerebrovascular reactivity (CVR). Several other sequences were collected during the scanning session related to other scientific questions and are not discussed further here.

The ME-MPRAGE sequence optimizes gray/white matter contrast ^24^ and covered the entire brain [1 mm isotropic voxels, 256 × 256 × 176 mm acquisition matrix, parallel imaging (GRAPPA) acceleration = 2, repetition time (TR) = 2,530 millisecond (ms), inversion time = 1,100 ms, flip angle (FA) = 7°, scan duration = 5.88 min] and had four echoes [first echo time (TE1) =1.69 ms, echo spacing (ΔTE = 1.86 ms)]. The 3D FLAIR sequence covered the entire brain (1 mm isotropic voxels, 256 × 256 × 176 acquisition matrix, TR = 5,000 ms, TE = 388 ms, inversion time = 1,800 ms, scan duration = 6.45 min). A high-resolution, flow compensated, multi-echo, 3D spoiled GRE sequence with eight echoes (TR/TE1/ΔTE/FA = 24ms/2.98ms/2.53ms/15°) was acquired and used to create QSM images using Ironsmith, our in-house developed software described elsewhere ^25^. The entire brain was covered [1.2 mm isotropic voxels, acquisition matrix = 224 × 224 × 144, parallel imaging (GRAPPA) acceleration = 2, and scan duration = 6.18 min]. The CVR sequence covered the entire brain [voxel size of 3.0 × 3.0 × 3.7 mm^3^, acquisition matrix = 64 × 64 × 36, parallel imaging (GRAPPA) acceleration = 2, TR = 2,000 ms, TE = 30 ms, FA = 71°, number of volumes = 216, and scan duration = 7.20 min].

### 3.3 ePVS Counting

We used a validated, visual rating method for quantification of region-specific ePVS burden developed via collaboration between multiple consortia and intended to standardize ePVS assessment across the field of cSVD research ^26^. As described in our previous work ^8^, the method involves manually counting ePVS on T1 images, with additional reference to T2 FLAIR images and susceptibility weighted images. Counts were performed in each hemisphere on a single, axial slice of T1 images, within four regions of interest (ROIs) that are known to have the greatest burden of ePVS ^5,15,16,26^: the centrum semiovale, 1 cm above the lateral ventricles; the basal ganglia, at the level of the columns of the fornix, including the head of the caudate and the putamen (Figure 1); the midbrain, at the level of the cerebral peduncles; the hippocampus, at the level of the midbrain.

**Figure 1:**
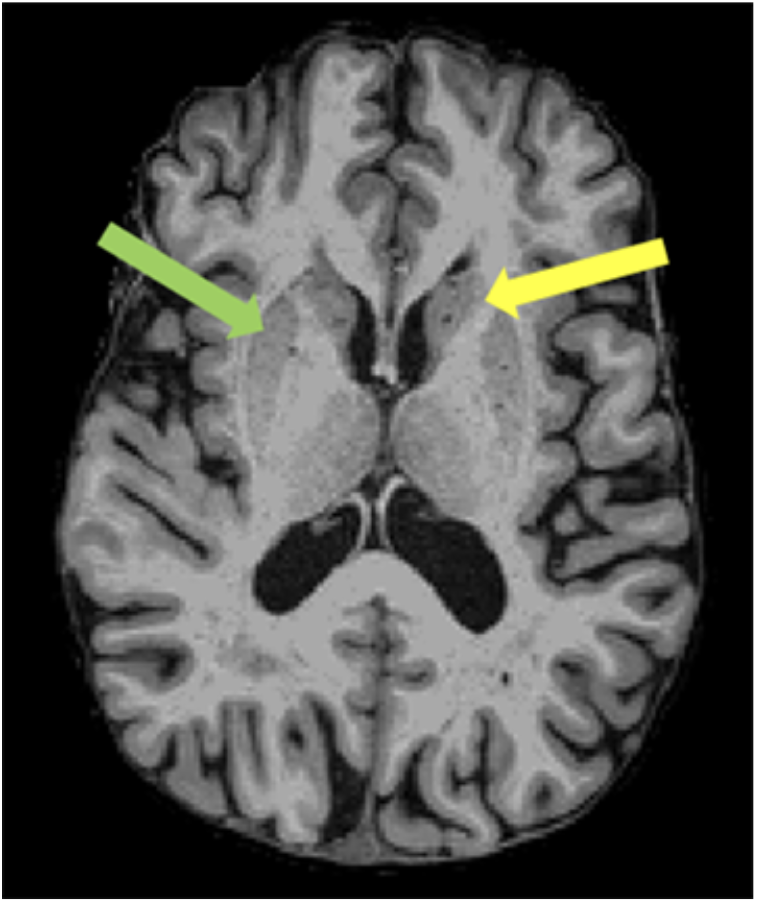
Example of basal ganglia ePVS on a T1-weighted image. An axial slice from representative participant is shown in the plane of the columns of the fornix. ePVS were counted within the head of the caudate (yellow arrow) and putamen (green arrow). A high intra-rater reliability (ICC = 0.9) was achieved for ePVS on a subset of 20 randomly selected participants.

Previous work by multiple consortia such as STandards for ReportIng Vascular changes on nEuroimaging (STRIVE) and Uniform Neuro-Imaging of Virchow-Robin Spaces Enlargement (UNIVRSE) have demonstrated the reliability of counting ePVS on T1, with very high correlation to counts on T2 images ^5,27,28^ as well as a high correlation between single-slice and multi-slice counts ^26,28^. All baseline and follow-up counts were conducted by the lead author (T.J.L.), blinded to participant demographics and neuropsychological scores, and under the supervision of an experienced neuroradiologist (F.D.R.), who clarified unclear imaging features.

In accordance with STRIVE and UNIVRSE consensus guidelines ^1,27,28^, ePVS were identified using T1, FLAIR and susceptibility weighted images. Prior to counting, each participant’s T1 images were registered to their FLAIR and QSM images in native space. ePVS were identified on T1 images as hypointense and less than 3mm in diameter to differentiate them from lacunes, which tend to be larger ^27,29^. ePVS were further differentiated from lacunes based on their lack of hyperintensity on FLAIR ^27,28^. ePVS were differentiated from cerebral microbleeds (CMBs) by their absence of prominent associated blooming artifact on QSM. We used QSM for differentiation of ePVS and CMBs due to evidence that QSM images outperform traditional single-echo susceptibility weighted images in this regard ^30^. Intra-rater reliability for ePVS was assessed on a subset of 20 randomly selected participants, using intra-class correlation coefficients (ICC).

### 3.4 Cerebrovascular Reactivity Imaging Procedure

CVR was assessed using a previously described procedure ^10,12,31^. Briefly, participants were fitted with a mouthpiece and nose-clip, and mild hypercapnic air (5% carbon dioxide, 74% nitrogen, and 21% oxygen) was administered using a Douglas bag, with a two-way non-rebreathing valve, enabling precise switching between room-air and hypercapnic air ^31^. Participants underwent blocked inhalation of hypercapnic air and room air while blood oxygen level–dependent (BOLD) MRIs were acquired continuously. A researcher was present inside the scanner room throughout the experiment to manually switch the valve to control the breathing of air (either room air or hypercapnic air from the Douglas bag). An interleaved, blocked fMRI design was used consisting of 3 blocks (50 sec/block) of hypercapnic air, and 4 blocks (70 sec/block) of room air for a total of 430 seconds. CO_2_ concentration in the exhaled air was sampled at 100 Hz and the resulting CO_2_ trace was recorded using capnography (Philips Respironics NM3 Monitor, Model 7900).

### 3.5 CVR Data Processing

CVR analysis was performed using a cloud-based online processing tool, CVR-MRICloud (Version 5, https://braingps.mricloud.org/cvr.v5) ^32,33^. Briefly, the BOLD data were first motion corrected and smoothed by an 8mm Gaussian kernel using SPM12. The end-tidal CO_2_ (Et-CO_2_) was extracted from the CO_2_ trace using an algorithm to identify the peak CO_2_ of each exhaled breath. The Et-CO_2_ curve was then temporally aligned with the whole-brain averaged BOLD signal time course. Whole-brain CVR values were subsequently obtained using a general linear model (GLM) in which whole-brain averaged BOLD signal was the dependent variable and temporally-aligned Et-CO_2_ was the independent variable. The CVR values were scaled to units of %BOLD signal change (in the CO_2_-enhanced vs. normal air condition) per mm of mercury (Hg) of Et-CO_2_ change (%BOLD/mmHg CO_2_).

Next, the BOLD images for individual participants were co-registered to the T1W ME-MPRAGE [(the four echoes averaged into a root mean square (RMS)] image. The T1W images were then segmented into ROIs by the T1 MultiAtlas Segmentation toolbox on MRICloud using the “Adult50-90yrs_287Labels30atlases_M2_252_V10A” atlas. These atlas-derived ROI masks were applied to the BOLD images to obtain ROI-averaged BOLD time courses, which were then used in GLM analyses similar to the whole-brain CVR calculation to obtain regional CVR values in specific ROIs (basal ganglia, hippocampus, and midbrain). An ROI mask of the centrum semiovale, which is not included in the CVR-MRICloud toolbox atlas, was also created via an in-house developed MATLAB script and FreeSurfer-based masks of the lateral ventricles. Specifically, the centrum semiovale ROI was defined as the portion of a FreeSurfer-derived, whole-brain white matter mask situated between two axial planes, relative to the lateral ventricles. The superior plane of the ROI was 12mm above the superior margin of the lateral ventricles and the inferior plane was 4mm above the superior margin of the lateral ventricles.

Lastly, CVR-MRICloud toolbox also provides the BOLD-CO_2_ correlation coefficient as a quality control measure. The BOLD-CO_2_ correlation coefficient was used to compare the relative coupling between BOLD signal and Et-CO_2_ between our four ROIs to assess potential differences in ROI signal quality.

### 3.6 White Matter Hyperintensity Quantification

Whole brain white matter hyperintensity (WMH) volumes were computed for use as a control variable in our models testing if CVR predicts change in ePVS after controlling for neuroimaging cSVD variables. Baseline total WMH volumes were computed using the UCD WMH segmentation toolkit (Version 1.3), which employs a validated 4-tissue segmentation method ^34^. This pipeline was chosen as it is also used for the Alzheimer’s Disease Neuroimaging Initiative (ADNI). Briefly, participants’ ME-MPRAGE image [the four echoes averaged into a root mean square (RMS) image] were first registered to their FLAIR image using FLIRT from FMRIB Software Library version 6.0.1 ^35^. The FLAIR image was then skull stripped, corrected for inhomogeneities using a previously published local histogram normalization ^36^, and then non-linearly aligned to a standard atlas ^34^. WMHs were estimated in standard space using Bayesian probability based on histogram fitting and prior probability maps. Voxels labeled as WMHs based on these maps exceeded 3.5 SDs above the mean WM signal intensity. Manual editing was performed by labeling false positive FLAIR hyperintensity as background. Total WMH volumes were calculated in participants’ native FLAIR space after back-transformation and reported in cubic millimeters.

### 3.7 Statistical Analyses

All statistical analyses were performed using SPSS (IBM, Armonk, NY, USA, version 28) with results considered statistically significant at p < 0.05. Independent sample t-tests and paired sample t-tests were used to examine differences between participants who were returners and non-returners as well as baseline and follow-up measures, respectively. To test the impact of baseline cerebrovascular dysfunction on longitudinal ePVS burden, a linear mixed effects model (restricted maximum likelihood, identity covariance structure) with random intercepts was performed. Specifically, the predictor variable was baseline CVR value in the basal ganglia and the dependent variable was follow-up basal ganglia ePVS counts after controlling for baseline basal ganglia ePVS counts. Time point was included as a factor in all models and additional covariates were age and sex. Planned models using CVR in other ROIs as a predictor were not run due to relatively low BOLD-CO_2_ correlation coefficient (as described in the Results section).

For the ROI showing a significant relationship between baseline CVR and longitudinal ePVS (i.e. the basal ganglia), two follow-up models were run to control for the potential influence of other relevant cSVD factors. In the first follow-up model, available participant-reported cSVD risk factors were added as covariates [body mass index (BMI), hypertension status, and type 2 diabetes status]. BMI was treated as a continuous variable and hypertension status and type 2 diabetes status were treated as dichotomous variables. In a second follow-up model, cSVD neuroimaging measures were added as additional covariates [whole brain WMH volume, lacune counts, and cerebral microbleed counts].

All predictors and dependent variables were tested for the assumption of normality using the Shapiro-Wilk test. Collinearity between predictors in all models was explored using the variance inflation factor (VIF), with a value of 5 implemented as a threshold value ^37^. Additionally, continuous covariates were centered.

### 3.8 Data Availability

The raw data supporting the conclusions of this article will be made available by the authors, without undue reservation.

## 4. Results

### 4.2 Data Characteristics

High intra-rater reliability was achieved for ePVS counts (ICC = 0.9) on a subset of 20 randomly selected participants. The mean follow-up time between the baseline and follow-up scan was 2.56 years (SD = 0.16 years). A summary of mean baseline, follow-up, and change in ePVS values in the 4 ROIs is presented in Table 2. Additionally, an illustrative case of ePVS progression during the follow-up period is presented in Figure 2. Of the 79 participants that underwent baseline MRI scanning, 50 returned for their follow-up MRI. This attrition rate is higher than is typical in our previous longitudinal studies with this cohort and likely reflects, at least in part, concerns related to participation in research during the COVID-19 pandemic. The use of linear mixed effects models, which better handle missing data, allowed us to minimize data loss and maximize power as compared to case-wise deletion methods. Returning participants were not significantly different than non-returners in age at baseline (t = 0.532, p = 0.596), sex (t = -1.093, p = 0.278), or ePVS counts in any of the four counting ROIs (All p > 0.05). However, returners had higher baseline MoCA scores (t = 2.148, p = 0.035) and showed a trend toward having higher mean levels of education (t = 1.971, p = 0.052) than non-returners. During the follow-up period, 7 of the 50 returner participants converted from cognitively normal to mild cognitive impairment based on their MoCA scores (range = 22-25).

**Table 2.** Summary of ePVS values in the 4 ROIs at baseline and follow-up

| ePVS ROI | Baseline<br>Count<br>(N = 79) | Follow-up<br>Count<br>(N = 50) | Mean<br>Increase<br>(N = 50) | %<br>Increase | t-<br>statistic | Two-sided<br>p-value |
| --- | --- | --- | --- | --- | --- | --- |
| Basal ganglia | 17.90<br>(4.54) | 21.32<br>(4.86) | 4.02<br>(2.73) | 23.2 | 10.415 | < 0.001*** |
| Centrum<br>semiovale | 52.13<br>(16.98) | 54.34<br>(15.15) | 4.88<br>(5.40) | 9.9 | 6.394 | < 0.001*** |
| Midbrain | 3.81 (2.67) | 4.72 (2.49) | 1.22<br>(2.10) | 34.9 | 4.103 | < 0.001*** |
| Hippocampus | 5.70 (2.79) | 5.98 (2.58) | 0.44<br>(2.49) | 7.9 | 1.248 | 0.218 |

This table lists the mean baseline ( ± sd), follow-up ( ± sd) and change ( ± sd) in ePVS across time points. Paired t-tests show ePVS significantly increased in the basal ganglia, centrum semiovale and midbrain, but not in the hippocampus.
\*\*\* $p \leq 0.001$

**Figure 2:**
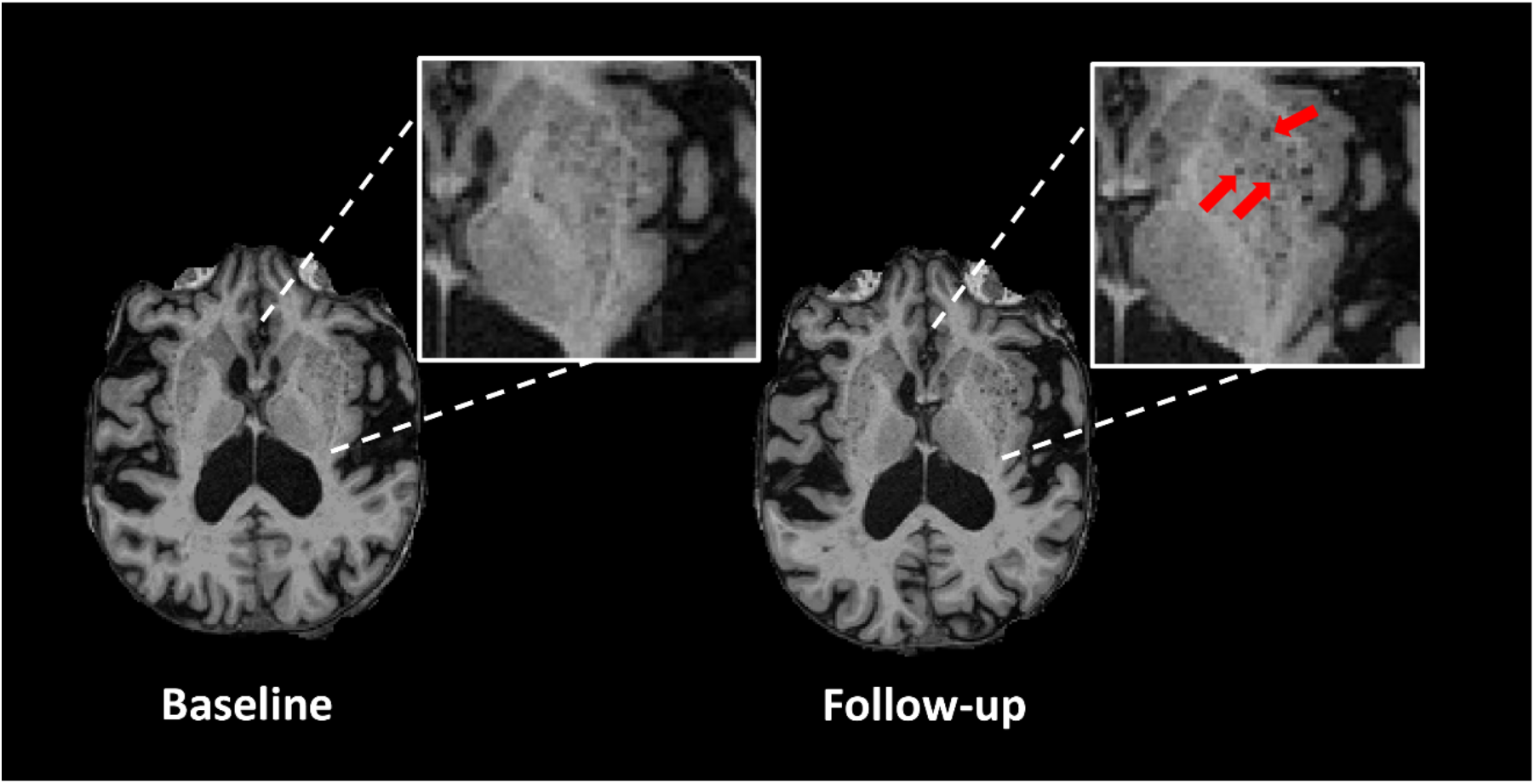
Example of basal ganglia ePVS burden at baseline and follow-up scan in a representative participant. Inset shows high ePVS burden and red arrows indicate several ePVS that developed during the follow-up period.

BOLD-CO_2_ correlation coefficient, a quality control measure of CVR provided by the CVR-MRICloud toolbox, was significantly higher in the basal ganglia than in the centrum semiovale (t = 73.3, p < 0.001, Supplementary Table 1), midbrain (t = 12.6, p < 0.001, Supplementary Table 1), and hippocampus (t = 10.3, p < 0.001, Supplementary Table 1). For this reason, CVR values in the centrum semiovale, midbrain, and hippocampus were not included in any additional analyses.

CVR values in the basal ganglia (W-statistic = 0.927; p < 0.001) as well as whole brain WMHs (W-statistic = 0.581; p < 0.001) were skewed and therefore log-transformed. Variance inflation factor for all predictors was < 2 and tolerance was > 0.5 in all analyses. Error residuals from all ePVS analyses were normally distributed indicating that the assumption of normality was met.

### 4.2 Relationship between Baseline CVR Values and Longitudinal ePVS Values

All results for associations between baseline CVR and ePVS counts are reported in Table 3. Baseline log-transformed CVR values in the basal ganglia significantly predicted an increase in basal ganglia ePVS counts over 2.5 years after controlling for baseline ePVS, age, and sex (coefficient estimate (SE) = -15.87 (3.92), p < 0.001, 95% confidence interval [CI] -23.68 to - 8.05). Baseline CVR values in the basal ganglia did not significantly predict increased ePVS in the centrum semiovale, hippocampus, or midbrain (Supplementary Table 2). To assess possible lateralization effects, the relationships between left and right basal ganglia CVR values and corresponding ePVS values were explored. Results indicated that lower baseline log-transformed CVR values in the left basal ganglia significantly predicted increased ePVS values in the left basal ganglia over time (coefficient estimate (SE) = -7.62 (4.78), p < 0.001, 95% CI -11.85 to -3.38). Similarly, lower baseline log-transformed CVR values in the right basal ganglia significantly predicted increased ePVS values in the right basal ganglia over time (coefficient estimate (SE) = - 7.91 (2.15), p < 0.001, 95% CI -12.19 to -3.62).

**Table 3.** Mixed effects model: relationship between log-transformed baseline CVR and longitudinal ePVS

| ROI | Model 1 | Model 2 | Model 3 |
| --- | --- | --- | --- |
| Basal Ganglia | -15.87 (3.92), $p < 0.001^{***}$, CI: -23.68 to -8.05 | -15.03 (4.00), $p < 0.001^{***}$, CI: -23.02 to -7.05 | -13.99 (4.03), $p = 0.001^{***}$, CI: -22.03 to -5.95 |
| Left Basal Ganglia | -7.62 (2.13), $p < 0.001^{***}$, CI: -11.85 to -3.38 | -7.32 (2.16), $p < 0.001^{***}$, CI: -11.62 to -3.01 | -6.62 (2.11), $p = 0.003^{**}$, CI: -10.84 to -2.40 |
| Right Basal Ganglia | -7.91 (2.15), $p < 0.001^{***}$, CI: -12.19 to -3.62 | -7.47 (2.20), $p = 0.001^{***}$, CI: -11.86 to -3.08 | -7.23 (2.22), $p = 0.002^{**}$, CI: -11.66 to -2.79 |

Values are measure coefficient estimate (SE), p value, 95% Confidence Interval. ePVS, enlarged perivascular spaces; BG, basal ganglia.
Model 1: adjusted for age, sex, and time point.
Model 2: adjusted for covariates in Model 1 and participant-reported hypertension status, type 2 diabetes status, and BMI
Model 3: adjusted for covariates in Model 2 and lacunes, cerebral microbleeds, and log-transformed whole brain WMH
\* $p < 0.05$
\*\* $p \leq 0.01$
\*\*\* $p \leq 0.001$

Two additional models were conducted to determine the impact of including additional cSVD covariates on the significance of the CVR-ePVS basal ganglia models. Model 2 accounted for participant-reported cSVD risk factors including hypertension status, type 2 diabetes status, and body mass index (BMI). In addition to the cSVD risk factors in Model 2, Model 3 further accounted for several known cSVD neuroimaging markers: lacunes, CMBs, and log-transformed whole brain WMH volume. Results indicated that CVR remained a significant predictor of increased ePVS counts in the basal ganglia over time after controlling for participant-reported cSVD risk factors (Model 2; Table 3) and additional cSVD neuroimaging markers (Model 3; Table 3). In addition, when testing potential lateralization effects, the relationships between individual left and right basal ganglia CVR values and corresponding longitudinal basal ganglia ePVS values remained significant after accounting for both participant-reported cSVD risk factors (Model 2) and additional cSVD neuroimaging marker covariates (Model 3).

## 5. Discussion

We tested whether baseline cerebrovascular reactivity (CVR) contributes to the later development of enlarged perivascular spaces (ePVS) in community-dwelling older adults. Our results showed that lower CVR values in the basal ganglia at baseline predicted a longitudinal increase in ePVS counts in the basal ganglia over a 2.5-year follow-up period. This finding supports a view that cerebrovascular dysfunction contributes to the development of ePVS in the basal ganglia.

Importantly, the relationship we observed between CVR and ePVS could not be accounted for by other commonly studied markers of cSVD. Specifically, the relationship between CVR and ePVS in the basal ganglia remained significant after controlling for the participant-reported cSVD risk factors of hypertension status, type 2 diabetes status and BMI (Model 2). In addition, the longitudinal relationship between CVR and ePVS in the basal ganglia remained significant after controlling for commonly used neuroimaging markers of cSVD including WMH volume, lacune count, and cerebral microbleed count (Model 3). Together, these results indicate that cerebrovascular compliance in the basal ganglia, as measured by BOLD-CVR, contributes to the subsequent development of ePVS in the basal ganglia.

One pathway through which reduced cerebrovascular compliance may lead to increased ePVS burden relates to glymphatic waste removal. The functional measure of cerebrovascular reactivity (BOLD-CVR) is an indicator of cerebrovascular compliance and reserve capacity ^11,38^. Diminished cerebrovascular compliance and reduced reserve capacity may impair vasomotion and pulsatility, which are required for effective glymphatic waste elimination ^3,9,39^. Reduced glymphatic waste removal may in turn lead to accumulation within the PVS and increased ePVS burden ^3^.

While our findings are novel, they are consistent with results from several studies that reported a relationship between non-neuroimaging measures of cerebrovascular dysfunction and PVS enlargement in the basal ganglia. For instance, basal ganglia ePVS are independently related to non-imaging measures of arterial stiffness ^17,18^, hypertensive arteriopathy ^19,40^, and atherosclerosis ^41^. Localization of such findings to the basal ganglia may be in part a result of an increased susceptibility to the early effects of cSVD in the perforating arterioles that supply the basal ganglia ^4,42^.

Our results further suggest that rates of ePVS accumulation may differ as a function of brain region. In our sample, a significant longitudinal increase in ePVS burden was observed in 3 brain regions (the basal ganglia, centrum semiovale, and midbrain). In contrast, the hippocampus showed the lowest rate of increase (7.9%), which was not significant in our sample. Future research with larger sample sizes will be needed to establish normative data on ePVS accumulation in different brain regions, the results of which will be of importance to the planning of future clinical trials that use ePVS as an outcome measure.

The main limitation of our study is that BOLD-fMRI is less sensitive in ROIs associated with fewer blood vessels. This resulted in comparably lower BOLD-CO_2_ correlation coefficient (quality control) values in the centrum semiovale, midbrain, and hippocampus, which prevented us from performing formal analyses in these ROIs. Furthermore, the relationship between BOLD signal and CO_2_ partial pressure may not be linear ^43^. As other human biomarker studies, our methods do not allow for conclusions about direct mechanisms. Future research using animal models will be required to delineate the mechanistic pathways through which poor vascular compliance contributes to ePVS development. An additional limitation to our study is our cohort included primarily highly educated, primarily White participants. Our findings will need to be replicated in more ethnically and demographically diverse cohorts.

### Conclusions

Our results indicate that cerebrovascular dysfunction is a contributing factor to ePVS accumulation in the basal ganglia. Future work should explore additional contributors to ePVS accumulation including the overproduction of proteins associated with neurodegeneration, including β-amyloid and tau. A better understanding of the mechanistic contributors to ePVS accumulation could point to early intervention targets intended to slow or prevent the symptoms associated with cSVD.

## Acknowledgments

We thank the dedicated research volunteers that make this work possible. We also thank Beverly Meacham, Eric Forman, Beatriz Rodolpho, and Stephen Dundon for assistance with MRI scanning, and Dr. David Powell for assistance with pulse sequence programming and selection. The authors declare no competing financial interests.

